# Mechanics of Contagion: A Newtonian Approach to Epidemiological Modeling

**DOI:** 10.1101/2025.08.13.25333606

**Authors:** Fernando Córdova-Lepe

## Abstract

**Resumen:** In this article, we establish foundational concepts, relationships, and principles to motivate and propose a “Mechanics of Contagion”, particularly its dynamic component. Methodologically, we employ analogical reasoning by constructing a simile within the context of the population-level spread of pathogens, drawing on the cause-and-effect relationships found in classical mechanics, a more widely accepted theoretical corpus. Specifically, we identify the causal mechanisms governing the physical motion of rigid bodies by examining the notion of forces, which explains variations in transmission rates within the flow from susceptible (S) to infectious (I) individuals in an SI-type mathematical model. Thus, it becomes meaningful, for example, to introduce concepts such as susceptible mass, infectious momentum, or contagion force. Finally, we formulate analogs of Newton’s three laws, demonstrating some applications to highlight the conceptual gains of the proposed Newtonian framework.

## 1 Introduction

In general, it is understood that classical mechanics (CM) is the branch of physics that studies the motion of macroscopic bodies by examining their causal factors (13). In this sense, it concerns itself with kinematics (the description of motion without considering its causes) and dynamics (the analysis of forces that produce or modify motion), as well as statics, which studies systems in equilibrium (i.e., absence of motion).

On the other hand, CM is likely the most universally recognized theory in physics due to its simple explanatory power and broad applicability (37), as well as its extensive metaphorical use for understanding other sciences (17; 30; 64). This is particularly true in epidemiology, at least in the context of certain conceptual frameworks (8; 67; 40).

In a general sense, motion is not merely physical; within abstract constructs, it can also be interpreted as change, both from certain philosophical perspectives (21; 38) and from more practical frameworks. In systems theory, motion is described as the temporal displacement or trajectory of a set of state variables within a phase space, characterizing a given system.

Note that, in epidemiological analysis, particularly from the perspective of mathematical epidemiology, contagion processes are viewed through a systemic lens. These systems, as mathematical objects, often take the form of differential or difference equation systems that resolve dynamics into trajectories (solutions) referred to as orbits (7; 9; 66).

Through an expanded conceptualization of motion, the primary objective of this work is to systematically apply the consolidated concepts, principles, relationships, and practical developments of Newtonian mechanics to the field of infectious disease epidemiology. This is achieved through methodical analogical reasoning (22; 6; 23; 42), specifically by postulating both formal and semantic analogies that allow the transposition of fundamental definitions and propositions from CM. Such an approach aims to establish sufficient foundational elements to formulate conjectures and, consequently, propose advancements toward a mechanical theory of contagion.

In this regard, previous work includes the kinematic component for the mechanics of contagion presented by Córdova-Lepe *et al*. (15). This article establishes epidemiological analogs for the physical concepts of position, displacement, and velocity. The analogy enables the presentation, within the context of a compartmental SIR model for a population of size *N* with basic reproductive number ℛ_0_, of the parameter **v**_0_ := ℛ0 − 1 as a (uniform) speed in a specific metric space (Ω, **d**). Here, Ω = (*S/N, I/N, R/N*) ∈ ℝ+^3^ : *S*+*I*+*R* = *N*, with the time counter being the variable *R* (the cumulative number of individuals removed).

Another relevant study (16), which aims to represent causality in variable transmission rates (*e*.*g*., as observed during pandemics), introduces a dynamic law (that is, one governing transmission rates) formulated as a spring-type differential equation. This work provides insights into contagion dynamics under the implementation of non-pharmaceutical mitigation measures and the decrease in population compliance with such strategies. In particular, it presents applications that successfully explain empirical data from the COVID-19 pandemic in Chile and Italy.

The present work extends beyond merely describing epidemic “motion”, instead aiming to explain its underlying causes through the development of fundamental epidemiological components and laws. This establishes a dynamic theory capable of supporting a comprehensive mechanics of contagion. Our approach employs conceptual analogs of momentum, impulse, force, and the governing laws of these quantities.

Of particular practical interest for theory building is our proposed (though not empirically proven) analog to Newton’s second law. This formulation would enable investigation of the consequences arising from various pressures that might be exerted on infection rates. While the literature contains some approaches applying physics concepts - particularly motion-related principles - to infectious disease epidemiology (*e*.*g*., in statistical mechanics (70; 33; 69)), our work systematically develops these connections.

In seeking to understand epidemic processes, our preferred approach favors mechanistic (white-box) models over phenomenological (black-box) ones. Mechanistic models tend to be explanatory, representing causal relationships through system structure and connections between variables, contingent on existing knowledge of the system (44; 31). In epidemiology, compartmental models (SI, SIR, SEIR, and their derivatives - collectively termed SIR-type models) exemplify this mechanistic approach. They partition populations and explain transfers (flows) between compartments using fundamental principles (*e*.*g*., balance equations, mass action laws, proportionality relations, or similar constructs).

Naturally, developing a mechanics of contagion requires that model derivations maintain counterparts in physics. We note that SIR-type models possess not only mechanical foundations but also strategic simplicity. Consequently, we seek a theoretical framework that supports models that capture the essential features of contagion processes (35; 14) - specifically those that explain dominant, visually identifiable patterns, such as the characteristic macroscopic shapes of the epidemiological curves of COVID-19. In other words, we prioritize strategic models that embrace simplicity principles (12; 26; 58), even at the cost of reduced data resolution.

Our objective is to propose a foundational theory that is not only mechanistic but also simple (average-based) and deterministic, consequently yielding models with these same characteristics. The incorporation of stochastic elements (subject to probability distributions) must await subsequent development, though we acknowledge their epidemiological significance, particularly when accounting for human behavior.

The paper begins with the background motivating the development of a mechanics of contagion in Section 2. Section 3 discusses the concept of force and examines what we consider to be the problematic use of the term “force of infection”. Section 4 presents and justifies the analogical methodology employed in our approach. Section 5 focuses on introducing the fundamental kinematic and dynamic concepts necessary for establishing the new theoretical framework. The proposed contagion dynamics is presented in Section 6, while Section 7 develops the epidemiological analogs of Newton’s three laws. Section 8 demonstrates two practical applications of our theoretical framework. Finally, Section 9 provides concluding remarks and a general discussion.

## 2 Background and motivations for a theory of contagion

From the COVID-19 pandemic experience, numerous mathematical epidemiologists have concluded that standard SIR-type models—with their assumption of constant transmission rates—fail to adequately explain empirical data unless incorporating extensive population compartmentalization. Recall that the basic SIR model partitions the affected population (of size *N*) into susceptible (S), infectious (I), and removed (R) individuals, relating their respective numbers through the equations:

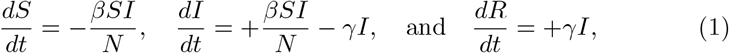

where *β* and *γ* are constant parameters representing the infection rate and recovery rate, respectively.

During the first year of the pandemic, before vaccine development and deployment, the system (1) and its derivatives proved insufficient to capture the characteristic geometric patterns observed in epidemiological curves across regional and national datasets (3; 57; 46). The key limiting factors appear to be human behavioral variability coupled with environmental fluctuations.

It is predictable that severe infectious diseases with high lethality will trigger diverse individual, social, and institutional responses. For instance, public health authorities and individual/collective awareness are expected to modify risk behaviors in disease transmission, whether by reducing the probability of human interactions or by enhancing pathogenic blocking mechanisms. Specifically, this involves increasing physical distancing, reducing mobility, or interrupting the potential transfer of pathogenic loads between hosts.

Within the framework of mathematical models, it is no longer tenable to assume that the transmission rate parameter (*β*, defined in (1)) remains constant over time, as human behavior is a dynamic factor altering transmission speed (see (59; 71)).

Thus, the transmission rate *β*(·) becomes a variable quantity, dependent on the average number of encounters an individual may have with others per unit time - a parameter highly sensitive to population-wide behavioral changes in response to perceived risk levels at any given moment. Numerically, the contact rate fluctuates according to variations in mobility patterns and social distancing practices. These variations arise both from implemented public health restrictions and from individual responses to such measures, which are intrinsically linked to the population’s risk perception.

Notably, in the context of COVID-19 and its basic compartmental models, analyses of different populations during the pre-peak epidemic phase revealed that estimating transmission rates as constant parameters proved unreliable, consistently generating case projections substantially higher than subsequently observed values. This leads to the critical conclusion that for explanatory modeling, key factors such as risk perception and population-level compliance with mitigation measures cannot be neglected (52; 68).

An increasingly critical scientific challenge lies in advancing our understanding of the dynamics governing the variable *β*(·)—specifically, explaining its value fluctuations across different epidemiological phases through both qualitative and quantitative causal analysis. Notably, transmission rate variability also stems from objective environmental conditions that either facilitate or inhibit pathogenic transmission.

Modelers have particularly focused on detecting and mathematically formalizing this variability. As emphasized in (29): “*Behavior is endogenous to a model when the parameter(s) associated with behavior is a function of another time-dependent variable within the model. Including behavior endogenously can enhance the utility of a model by providing a mechanism for how behavior varies in response to both control measures as well as the epidemic dynamics*.”

This perspective is further supported by (43; 55), who adopt similar theoretical approaches to behavioral incorporation in epidemiological modeling.

Recognizing that *β*(·) represents a transmission rate and that the change Δ*β* := *β*(*t*+Δ*t*) − *β*(*t*) over a time interval [*t, t*+Δ*t*] defines Δ*β/*Δ*t* as the rate of change of the transmission rate (rate of a rate), we observe that epidemiological phenomena invariably involve acceleration or deceleration of the dynamics of contagion. This leads us to investigate the causal mechanisms governing Δ*β/*Δ*t*, which we might call the acceleration of disease transmission.

Our motivation stems from establishing a conceptual bridge between epidemiological phenomena and physical forces as dynamic factors (whether internal or external) that either catalyze or inhibit transmission processes. Specifically, we propose to formulate, through analogy with CM, a notion of force associated with transmission acceleration. This theoretical construct would serve as the foundation for explaining both the kinematic and dynamic aspects of infectious disease epidemiology, positioning force as a fundamental concept in developing a comprehensive mechanics of contagion.

## 3 Reexamining ‘force of infection’: From Aristotelian legacy to Newtonian foundations

Aristotle, whose qualitative physics dominated for two millennia, explained motion through attraction/repulsion between the four elements (water, fire, earth, air). His concept of movement extended beyond spatial displacement to include qualitative changes (39). He distinguished between natural motions and violent motions, the latter requiring an external force as an efficient cause (21). While he never developed a population theory of contagion, Aristotle’s History of Animals (5) recognized that rabies was transmitted through bites from infected dogs. This observation, though pre-scientific by modern standards, established an early conceptual link between physical contact and disease transmission.

As the founder of mechanics (qualitative), Aristotle would have asserted that heavier objects require more force to set them in motion and that objects pushed with greater force move faster. This view, if expressed proportionally, leads to what we shall call the *Aristotelian force*:

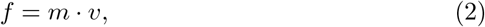

where *f* represents force, *m* denotes a body’s mass, and *v* its speed. This formula - a well-known example of a “misconception” in understanding natural phenomena - was ultimately superseded by Newtonian physics (34; 1).

In epidemiology, the term “force” appears in the expression *force of infection*, a concept that Ronald A. Ross reportedly introduced (as the per capita rate of acquisition of infection through infectious mosquito bites) (60). The Nobel laureate’s mathematical framework for malaria transmission theory established him as a foundational figure in epidemiological analysis. However, according to (33), the concept predated Hugo Muench’s 1934 work (47), where it was referred to as the *effective contact model*. The current terminology only became standardized after 1959 (48).

This force concept, when operationalized with empirical data, is directly expressed as the rate at which susceptible individuals within a given population become infected. More precisely, it is quantified through the ratio of new infections to the number of exposed susceptible during a specific exposure period. As can be inferred, the force of infection incorporates factors such as: population susceptibility, disease prevalence, and pathogen transmissibility. Note that as a rate measure, this indicator has units of time^−1^.

In what follows, for SIR-type models, we denote by the italicized variables *S, I*, and *R* both the epidemiological type and the corresponding group (set) and its size; the same applies to the total population *N*. Note that, in light of the structure of the Aristotelian force (2), the force of infection in some SIR-type models, such as (1), takes the form:

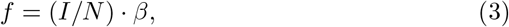

where *I/N* represents the fraction of infected biomass (as a proportion of individuals) in a population of size *N*. This corresponds directly to Definition I in Newton’s Principia (50): “*The quantity of matter is the measure of the same, arising from its density and bulk conjunctly*.” On the other hand, *β* denotes the transmission rate, which measures the infection speed. In particular, this Muench force of infection is expressed in units of time^−1^, contradicting the expected dimensions of an acceleration.

That velocity could increase at a constant rate (gravity) was Galileo’s seminal discovery through his experiments with spherical balls rolling on inclined planes. Perhaps most revolutionary was his expression of this principle in mathematical language. This foundational work enabled Newton to develop the concept of acceleration - a fundamental construct for establishing mechanics and the related concepts of momentum, force, and impulse as cornerstones for his laws of motion. The most significant of these states that “the net force applied to an object equals the product of its mass and acceleration,” succinctly expressed as:

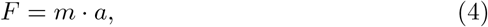

where *a* represents acceleration, defined as *dv/dt* (with *v* denoting speed). This constitutes Newton’s second law of motion.

The key conclusion is that the epidemiological concept of force of infection represents an analog to physical force, but in an Aristotelian sense. Now, recognizing that the infection rate is variable – meaning its variation constitutes an acceleration – we argue it is timely to formulate a Newtonian-inspired definition of infection force. This approach would enable us to reconceptualize contagion processes through classical mechanics analogies, thereby transferring much of its explanatory and predictive potential to epidemiological analysis.

This aspiration is by no means novel, as Newtonian mechanics, being a conceptual framework, has permeated numerous fields of knowledge, particularly those with strong mathematical foundations. Traditionally, these concepts have been most prominently applied in mechanical engineering, aerospace engineering, and astronomy, given their direct connection to physics as applied disciplines. However, other domains that were previously less expected to incorporate mechanical principles have progressively adopted them to explain dynamic systems. A notable example is biomechanics (the study of human and animal movement), which employs these concepts to understand the forces acting on biological systems’ physical components. Consequently, in applied fields such as ergonomics and prosthetic design, mechanical principles have become indispensable.

However, in domains where the objects of study are less tangible and require mental constructs, mechanics has also gained increasing relevance. This is the case, *e*.*g*., in econophysics within economics, where concepts from CM have been employed to explain the dynamic behavior of complex economic systems.

In other words, the challenge of developing a mechanics of contagion appears plausible, and this work aims to establish the foundations for such a possibility. In mathematical epidemiology, models are formulated through differential equations whose solutions—dynamic trajectories—closely resemble the equations of motion in physics. This suggests that establishing connections between epidemiological variables/parameters and mechanical concepts of forces and rates of change appears feasible, provided appropriate analogies can be drawn, particularly at the conceptual equivalence level.

We observe that the diffusion concept, fundamental to fluid mechanics and heat transfer, has significant applications in the study of the spatial spread of infectious diseases. By drawing analogies between populations and large-particle systems, elements of statistical mechanics could potentially be employed in epidemiological analysis. Another example emerges in control theory, where the principles of CM inform the design of mitigation strategies, for example, by identifying forces capable of altering transmission dynamics.

## 4 On the Analogical Method

Analogical reasoning—alongside deduction, induction, and abduction—constitutes a fundamental pathway for knowledge construction, particularly through modeling approaches (2; 63; 51). When formalized as an analogical method, this reasoning becomes particularly valuable for addressing complex processes, where we can identify parallel contexts that reveal conceptual similarities. These parallels enable us to establish conceptual relationships and patterns, thereby allowing us to infer characteristics and behaviors in complex systems based on better-understood frameworks.

Moreover, the advantages of analogical reasoning in knowledge acquisition are well-documented and extensively explored (28; 65).

Analogies serve as powerful tools for understanding complex processes by enabling the formulation of informed assumptions (45), particularly when conventional approaches are inadequate. They provide practical scaffolding, allowing researchers to explore unfamiliar domains through established scientific knowledge from analogous contexts. This article aims to establish foundational principles and outline implications for a mechanics of contagion, drawing upon the well-established framework of CM.

Within public health and epidemiology specifically, analogical reasoning has played an illuminating role in addressing knowledge gaps, as demonstrated in (19; 20; 27). Particularly noteworthy is its application in scientific concept formation (49; 23).

There exist notable cases of analogical constructions from epidemiology to other scientific disciplines, highlighting the bidirectional utility of this methodological approach (25). For instance, wildfire propagation processes have been conceptualized through epidemiological frameworks, where fire is represented as a pathogenic agent triggering outbreaks (62; 10; 56; 4). Currently, we observe the study of viral propagation in computational systems through direct conceptual and model-to-model parallels with epidemiological terminology from biological systems (72; 53; 18). Analogical reasoning constitutes a scientific practice that enhances understanding and enriches conceptual meanings (24; 36).

We note that analogical methods may involve either semantic analogies (focusing on the conceptual meaning of relationships) or so-called syntactic analogies (concerned with structural or formal similarities). In this work, we employ both approaches, conceptual definitions and mathematical formulations, to introduce the notion of contagion force, that is, to address both the meaning and syntax of equations. This dual approach mirrors the analogous relationship between formulations in gravitational mechanics and classical electromagnetism (32).

## 5 Concepts for a mechanics of contagion

In what follows, to present the theory within a simplified framework, in (1) we assume that *γ* = 0, *i*.*e*., we consider the *β*(·)-SI model (an SI model with variable transmission rate). In its simplest form, this model assumes a constant population size *N*, giving the following:

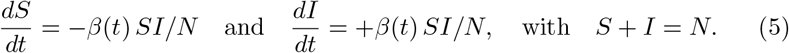

Regarding the challenge of population-scale mechanics of contagion processes, we must assume, in analogy with CM, the existence of a kinematic and a dynamic perspective; that is, we must focus on both the description and explanation of movements between the epidemiological classes involved, primarily concerning the flow from the susceptible group to the infected. Among the concepts of Newtonian mechanics, somehow present in a mechanics of contagion, are those of kinematics combined with those of dynamics: mass, momentum, force, and impulse.

### 5.1 Kinematics: position, displacement, and velocity

Kinematics addresses the movement of individuals between epidemiological states, where for the system (5), the change corresponds specifically to transitions from susceptible to infectious status. The description of such movement fundamentally requires the concepts of space and at least one mobile unit. In this framework, the mobile unit is represented by the time-dependent point (*S*(·), *I*(·)) within the bounded linear space Ω := (*x, y*) : *x, y* ≥ 0 and *x* + *y* = *N*. Core descriptive concepts include position, displacement, velocity (or speed), and acceleration.

This formulation allows analogizing the contagion process to the dynamics of a particle with mass *N* (population mass) whose trajectory (*S*(·), *I*(·)) evolves through the epidemiological mass space Ω. Therefore, the population position at any time *t* is uniquely determined by the coordinates (*S*(*t*), *I*(*t*)) in Ω-space. Note that the *displacement* corresponds to the change in position within the space. Between instants *t*_1_ and *t*_2_, *t*_1_ *< t*_2_, the position of the population has changed; the displacement is defined by the difference (*S*(*t*_2_) − *S*(*t*_1_), *I*(*t*_2_) − *I*(*t*_1_)), a vector lying in Ω. Furthermore, over each interval [*t*_1_, *t*_2_], the population’s position defines a curve in Ω. Considering that the trajectory of a particle is the locus of its successive occupied positions, it follows that the *epidemiological trajectory* of the entire population is the graph in Ω of the curve (*S*(*t*), *I*(*t*)), with *t* ∈ [0, ∞). Now, considering the vector of logarithmic derivatives under the 1-norm (sum of the absolute values of the coordinates), we have:

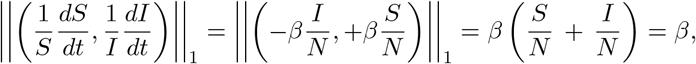

hence *β*, whether constant or variable, represents the relative velocity of the system. When *β*(·) ≡ *β*_0_, *i*.*e*., a constant transmission rate, we designate (*S*(·), *I*(·)) as a *uniform epidemiological trajectory*, which we associate with inertia – indicating the absence of deviating forces.

### 5.2 Dynamics: mass, momentum, and force

The physical representation of a body corresponds to an extension unfolding in space, to which we associate attributes such as mass, shape, and motion. In the contagion dynamics of the *β*-SI model, we identify the coexistence of two distinct bodies: one constituted by the susceptible population and the other by infectious individuals.

#### 5.2.1 On a mass epidemiological concept

Regarding quantity, Newton (50) refers to mass as the quantity of matter, defining it as follows: “*The quantity of matter is the measure of the same, arising from its density and bulk conjunctly*”. Here, we observe the necessity for this definition of quantity of matter to remain invariant under changes of position or shape.

Now, returning to our analogy, consider a population of size *N* partitioned into subsets (or *types*) *X* ∈ {*S, I*}. For any subgroup *G* of the population, we define its *X-mass* and *X-density* respectively as:

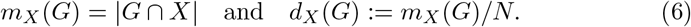

Thus, the *X*-mass of *G* represents the number of individuals in state *X*, while the *X*-density is the ratio of this mass to the total population size.

Specifically, for the infectious group (*X* = *I*), the infectious mass and density of *G* are given by:

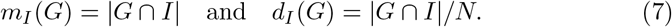

When *G* = *X* (*i*.*e*., when *G* coincides with either the susceptible or infectious compartment), we obtain: *m*_*X*_ (*X*) = |*X*| and *d*_*X*_ (*X*) = |*X*|*/N*. For notational simplicity, we denote these quantities by *m*_*X*_ and *d*_*X*_, respectively.

#### 5.2.2 On the quantity of movement concept

In Definition II of his *Principia*, (50), Newton states: “*The quantity of motion is the measure of the same, arising from the velocity and quantity of matter conjunctly*”. Therefore, for a mechanical theory of contagion, we must first clarify what we mean by the motion of the bodies *S* and *I*. In this regard, the most direct approach is to define motion as a change in the state of said body. In this sense, when a group of infectious individuals meets (contacts) a susceptible one, the latter ceases to be the same, as some of its components (individuals) changed their condition - if they were previously free from carrying the pathogen, they no longer are, becoming new hosts. Thus, the direct interest lies in quantifying the magnitude of this change.

From physics, we learned the difference in the magnitude of effort required to stop a train moving at 80 km/h compared to a motorcycle at the same speed, as these scenarios involve distinct *quantities of motion*. In an epidemiological context, given the same transmission rate *β*, the scenario where 10% of the population is infected differs fundamentally from one with only 1% prevalence. Both empirical evidence and intuition suggest that halting the infectious process is easier for the smaller group, even when both groups transmit at the same rate *β*.

This analogy will be pivotal for constructing a notion of *epidemic momentum*.

#### 5.2.3 On the concept of force

Within a specific thematic context, if we inquire about the properties a variable must possess to be termed a *force*, we must establish minimal conditions. In this regard:

(a) Its action, exerted by one object upon another, must alter the second object’s state of “motion” by accelerating it (positively or negatively), *i*.*e*., changing its rate and diverting it from uniform trajectory. Thus, we must define what constitutes a uniform trajectory for (*S*(·), *I*(·)). The natural case corresponds to a constant transmission rate *β*.
(b) It must be a numerically quantifiable variable (or its derivative), *i*.*e*., the force must be measurable. Furthermore, its units should reflect the analogy, implying units of count per inverse time squared. In our framework, where changes are measured in individuals (ind), we should have [ind/days^2^] if time is measured in days.
(c) When applied to an object over time, there must be an observable interaction effect. In our analogy, this effect manifests as changes in the *S* → *I* flow magnitude relative to the uniform scenario, *i*.*e*., through increases or decreases in *β* depending on the force’s direction.

## 6 A mechanics of contagion (its dynamics)

### 6.1 On the transmission rate

Theoretically, the transmission rate *β* represents the number of contacts per unit time that a specific individual Q may have with any other individual P, such that if prior to contact Q ∈ *S* and P ∈ *I*, then following contact we will certainly have Q ∈ *I*.

Thus, we observe that *β* incorporates two conjunctive components:

(a) a contact frequency, and
(b) the probability that under given epidemic conditions, a contact pair will produce a newly infectious individual.

Regarding the counting of contacts between individuals, to better understand this concept, we will define what we shall call an *epi-band*. Consider a generic individual P in the population who serves as the “marker” for a certain spatiotemporal band *V*_*δ,τ*_ (P), where *τ* (*τ <* 1) represents the minimum duration of spatial proximity within a maximum distance *δ* from P.

Given another individual Q, we say that Q had contact with P if there exists a non-punctual time interval *J* such that (*Q*, |*J* |) belongs to *V*_*δ,τ*_ (P). That is, if **d**(*Q, P*) *< δ* (for some distance metric **d**) and |*J* | *> τ*. Then,

- If an individual Q remains consistently beyond distance *δ* from P, then Q has not entered the epi-band, *i*.*e*., this does not count as a contact.
- If an individual was within distance *δ* of P, but remained so for a duration shorter than *τ* (throughout the observation time unit), this is not considered a contact.

The mental image is that an individual Q does not qualify as a contact of P if it remains distant from P (beyond *δ*), or stays nearby but for insufficient time (continuously for less than *τ*).

The specification of *V*_*δ,τ*_ (·) admits various geometric configurations depending on the dimensions of the parameters *δ* (spatial) and *τ* (temporal). While suggesting that a larger *δ* (*resp*. smaller) could be compensated by a smaller *τ* (*resp*. larger) if one aims to maintain an unchanged contact count.

We identify the types of contact numbers (CN), per unit time, presented in Table 1.

**Table 1:** Types of contact numbers (CN) - general, specific, or positive - within an epi-band *V*_*δ,τ*_ (·). Note that *κ*_*esp*_ = *κ/N* and *κ*_*inf*_ = *κ*_*esp*_ · *I* = *κ*_*gen*_ · (*I/N*).

| Sign | CN | Def.: Number of times per unit time that... |
| --- | --- | --- |
| $\kappa_{gen}$ | General | an individual $Q$ enters $V_{\delta,\tau}(P)$, for some $P$, $P \neq Q$. |
| $\kappa_{spec}$ | Specific | an individual $Q$ enters $V_{\delta,\tau}(P)$, given certain $P$, $P \neq Q$. |
| $\kappa_{inf}$ | Positive | an individual $Q$ enters $V_{\delta,\tau}(P)$, some $P$ in $I$, $P \neq Q$. |

However, the mere occurrence of contact between individuals from different epidemic groups, one susceptible and the other infectious, does not guarantee that the susceptible individual will transition into the infectious compartment. Under specific pathogen-dependent conditions and environmental contexts, we define *transmissibility* as the probability *ρ* that, given a contact between Q and P under the described conditions, a new infectious case is generated.

Note that the classical (Aristotelian) infection force *f* (see (3)) corresponds to what we may call the *effective contact number*, denoted *κ*_eff_. This represents the number of positive contacts where transmission is effectively expressed when occurring between individuals of crossed epidemic types. Therefore,

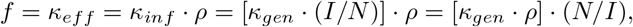

whereby

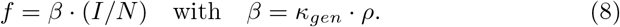

Thus, *β*, the transmission rate, within the mathematical formalism, represents a fraction of the general contact number – specifically, those contacts that are potentially effective.

This well-known equality is particularly interesting, as it decomposes the transmission rate *β* into: (a) a socio-behavioral component (*κ*_*gen*_), and (b) a pathogen-specific factor (*ρ*) accounting for its emission and environmental persistence.

Consequently, epidemic control strategies typically aim to reduce *β* through either the pathway in (8): decreasing contact frequency or blocking pathogen transmission.

A key consideration in our approach is the potential dependence of *β* on the epi-band *V*_*δ,τ*_ (·). Nevertheless, observe that narrower (*resp*. wider) neighborhoods unambiguously produce fewer (*resp*. more) general contacts *κ*_*gen*_ while simultaneously increasing (*resp*. decreasing) the transmission probability *ρ*.

### 6.2 Susceptibility and infectivity risks as epidemic momenta

In physics, momentum serves as an indicator of how difficult it is to alter the velocity of a moving object, defined as the product of an object’s mass and its velocity. The natural analogous question would be: how challenging is it to halt a contagion process?

Susceptibility alone does not provide a clear measure of risk for the development of a contagion process within a population. Indeed, without a transmission rate, no risk would exist. Thus, the risk cannot depend solely on the size of the susceptible subpopulation, given that this population exhibits a time-varying transmission rate at each instant. A similar observation applies to infectiousness. Therefore, we define:

- The *S*-*momentum* or *susceptibility risk*, defined as **P**_*S*_ := *S* · *β*.
- The *I*-*momentum* or *infectivity risk*, defined as **P**_*I*_ := *I* · *β*.

Note that the units of **P**_*X*_ (*X* ∈ {*S, I}*) are [individuals/time].

Thus, as expected, the susceptibility risk per susceptible individual and the infectivity risk per infectious individual both equal the transmission rate:

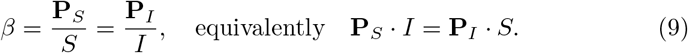

It is also possible to refer to the susceptibility or infectiousness of a subgroup within the population. If *G* is a group of individuals in the population, we can define the *susceptibility risk of G* as **P**_*S*_(*G*) := *m*_*S*_(*G*) · *β* and the *infectivity risk of G* as **P**_*I*_ (*G*) := *m*_*I*_ (*G*) · *β*.

### 6.3 Impulse formulation and time-averaged contagion forces

The concept of impulse over a time interval [*t*_1_, *t*_2_], in physics, corresponds to the change in momentum. Thus, we would have an impulse associated with this interval for both susceptible and infectious individuals, given by:

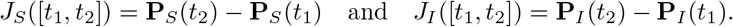

When *β*(·) is constant (*β* ≡ *β*_0_), denoting Δ*X* = *X*(*t*_2_) − *X*(*t*_1_) for *X* ∈ {*S, I}*, we obtain:

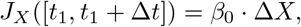

which has units of [individuals/time].

Note that

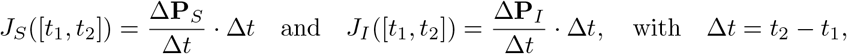

inspire the definitions of the *mean susceptible contagion force* and *mean infectious contagion force*, respectively, as:

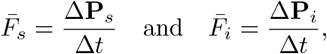

with units of [individuals/time^2^].

### 6.4 Instantaneous contagion forces: Susceptible and infectious formulations

In classical mechanics, force is what, when applied to a body, can alter its state of motion. One possible state is uniform evolution, which, according to the previously developed kinematics, corresponds to a constant transmission rate (*i*.*e*., non-accelerated). Now, if *β* (a velocity analog) changes, acceleration occurs. Where there is acceleration, we may then speak of the presence of a force.

The following definition, by direct semantic analogy, is that of the *susceptible contagion force* (or *infectious contagion force*), which as an instantaneous force represents the limit process of their mean versions. Thus, we have:

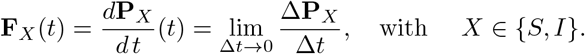

Thus, the instantaneous force is the derivative of the product *β*(·) *X*(·), where *X* ∈ {*S, I}*.

For the case *X* = *I*, we obtain:

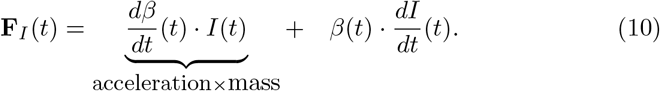

In (10), the first term is analogous to (4), while the second accounts for the case of non-constant mass.

Now, considering that *dI/dt* = *β*(*t*) · *S* · (*I/N*) and that *β*(·) = *κ*_gen_(·) *ρ*(·), while omitting time arguments for clarity, we obtain:

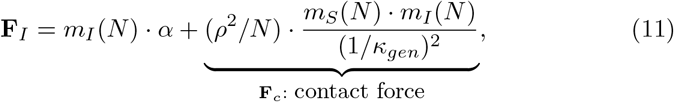

where *α* := *dβ/dt*, which—being an acceleration—we may term the *transmission rate change rate*. Note the clear syntactic analogy (in form) between the term we have called the *contact force* and Newton’s law of gravitation, which would be the sole term if the rate *β* were constant. Thus, in what we have called the uniform case, a force is always acting.

Observe that for a given epi-band, populations exhibiting a higher (*resp*. lower) general contact number *κ* inherently maintain less (*resp*. more) social distancing. Thus, it is not counterfactual to denote 1*/κ* as the *epi-distance*. Regarding the *ρ*^2^*/N* factor, this is not a universal constant like the gravitational constant, since we know *ρ* depends on both the epi-band and the medium’s permittivity for pathogen jumps to new hosts. In this sense, the analogy aligns more closely with the constant in Coulomb’s Law of electromagnetism.

To better understand the dependence of **F**_*c*_ in (11) on population size, we can express it using normalized susceptible and infectious masses. In this case:

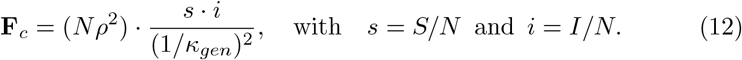

We need only observe that for the infectious contagion force, the calculations are similar. More precisely:

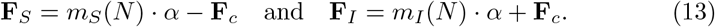

## 7 Epidemiological analogues to Newton’s Laws

For a system of interacting entities, a “force” (in a broad, operational sense) is the interaction that provides a causal explanation for inducing changes in their motion (velocity). The epistemological approach of understanding through forces is rooted in prior knowledge; consequently, we may expect historical but primarily pragmatic reasons for this framework. In this context, Newton’s Laws have established causality while enabling mathematical formalization, making their metaphorical transposition to epidemiology inevitable.

### 7.1 First Law (Inertia)

> “*Every body continues in its state of rest, or uniform motion in a right line, un less it is compelled to change that state by forces impressed upon it*” (50).

We are clear that inertia requires the absence of acceleration, so in (13) we set *α* = 0, that is, we reduce the contagion force (whether susceptible or infectious) solely to the contact force **F**_*c*_. Thus, trajectories under uniform transmission (*β* constant) are governed exclusively by **F**_*c*_.

How then do we link this case to the concept of inertia? The answer lies in the observation that when two susceptible population groups encounter (contact) the same infectivity risk, they will yield identical per-unit-time counts of individuals transitioning from susceptible to infectious states.

***1***.^*st*^ **Law of contagion**: “*Every population remains in a state of infection-free equilibrium or uniform epidemiological trajectory, unless compelled to change by an external contagion force*.”

### 7.2 Second Law (Dynamics)

> “*The change of motion is proportional to the motive force impressed; and is made in the direction of the right line in which that force is impressed*.” (50).

The force exerted by a body of infectious individuals on a susceptible one is mediated through interpersonal proximity—that is, via close contacts. However, the mechanisms enabling this proximity may vary, *e*.*g*., through human behavior (altering *κ*_eff_) or environmental/pathogen-intrinsic factors (modifying *ρ*), thereby varying the transmission rate.

The analogous conjecture is that the excess infectious contagion force (**F**_*I*_ − **F**_*c*_) equals the sum of external forces acting on the system:

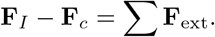

***2***.^*nd*^ **Low of contagion**: *The rate at which the risk of infectivity (or susceptibility) changes is equal to the contact force plus the net force (sum of all forces) acting on a body of infectious individuals (or susceptible individuals)*.

### 7.3 Third Law (Action and Reaction)

> “*To every action there is always opposed an equal reaction: or, the mutual actions of two bodies upon each other are always equal, an directed to contrary parts*.” (50).

From the equation (−*βm*(*S*)*I*)+(+*βm*(*I*)*S*) = 0 emerges a momentum principle **P**_*S*_*/S* = **P**_*I*_*/I*, which must have a force counterpart expressed through the action-reaction principle. Note that in (13), summing **F**_*S*_ and **F**_*I*_ yields **F**_*S*_ + **F**_*I*_ = (*S* + *I*) · *α*, leading to **F**_*S*_ = −**F**_*I*_ + *N* · *α*, which simplifies to **F**_*S*_ = −**F**_*I*_ when *β* is constant, as expected.

***3***.^*rd*^ **Low of contagion**: *To every action on susceptibles there corresponds an equal action on infectious individuals - provided the transmission rate remains constant*.

## 8 Some applications

### 8.1 A *β*(·)-SI model without external forces

This case should correspond to the uniform situation. Indeed, if **F**_ext_ = 0, the second law of contagion becomes **F**_*I*_ − **F**_*c*_ = 0. Using (13), we obtain *m*_*I*_ · *α* = 0, so that *α* = 0, and consequently *β* is constant.

### 8.2 A *β*(·)-SI model with mitigation force

Note that during an active transmission process, health authorities implementing mitigation measures may aim to reduce the transmission rate. Within our *β*-SI framework, they could apply an external force proportional to both the daily new case count and the current infection rate value, *i*.*e*.,

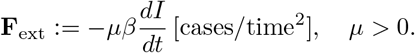

So, according to the postulated *second law of contagion*, this must balance the infection force against the contact force: **F**_*I*_ (*t*) − **F**_*c*_ = **F**_ext_(*t*), which leads to *{dβ/dt}I* = −*µβ*(*t*)*{dI/dt}*. This implies:

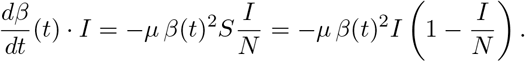

Thus, assuming *I* ≠ 0, the *β*-*SI* model (*i*.*e*., with variable transmission rate) in terms of *i* = *I/N* is defined by the following differential system:

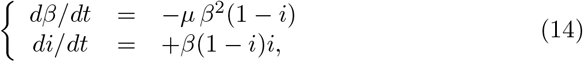

with (*β, i*) ∈ (0, ∞) *×* (0, 1].

Note that it is clear that *i*(·) is strictly increasing and converges to one at infinite time. On the other hand, by isolating (1 − *i*) from both equations in (14), we obtain (*dβ*(*dt*)*/β* + *µ* (*di/dt*)*/i* = 0. Therefore, for an initial condition (*β*_0_, *i*_0_) at time zero, integration yields:

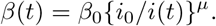

Since *i*(·) is increasing with *i*(∞) = 1, we have that *β*(·) decreases monotonically from *β*_0_ toward 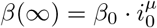.

### 8.3 A *β*(·)-SI model with mitigation and contravention

In the previous *β*(·)-SI model, in addition to the external mitigation force **F**_ext(1)_ = −*µβ*(*t*)(*dI/dt*), there exists a second external force **F**_ext(2)_ of non- compliance (population non-adherence) related to the existence of an intrinsic transmission rate *β*^*^, toward which the system tends to return. This behavioral external force drives the rate upward (toward the intrinsic value) and is proportional to: (1) the gap *β*^*^ − *β*(·), (2) the infectious load, and (3) the number of susceptibles, *i*.*e*.,

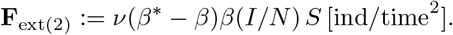

**Figure 1.**
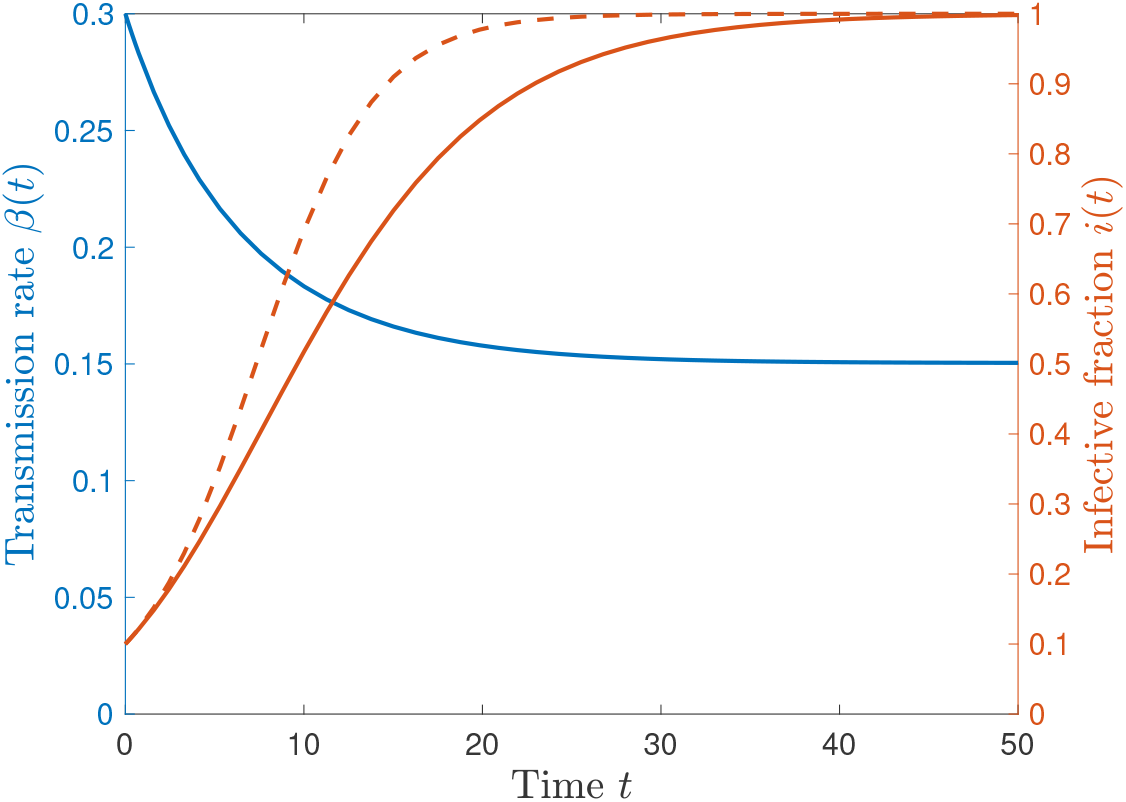
Temporal dynamics of the transmission rate *β*(*t*) and infected fraction *i*(*t*) according to model (14), with trajectories represented by the blue and red curves, respectively. Additionally, the red dashed line represents the trajectory of the classical SI model. The parameter is *µ* = 0.3, and initial conditions are *β*_0_ = 0.3 and *i*_0_ = 0.1, from which the asymptotic value *β*(∞) = 0.1504 is obtained..

This constitutes a positive force, as it acts in the same direction as the infectious variable.

As before, according to the second law of contagion, we equate the force difference **F**_*I*_ (*t*) − **F**_*c*_ with **F**_ext(1)_(*t*) + **F**_ext(2)_, which leads us to the equation:

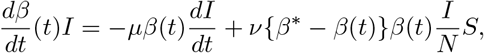

whereby

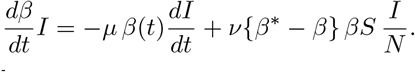

This is, if *I*≠ 0, we have

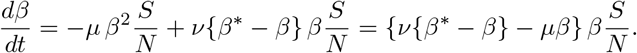

Thus, the *β*-*SI* model with variable transmission rate, expressed in terms of *i* = *I/N*, is defined by the following differential system:

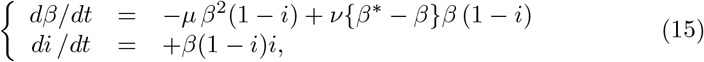

with (*β, i*) ∈ (0, ∞) *×* (0, 1].

Note that by defining *λ* = *µ* + *ν* and isolating *β* (1 − *i*) from both equations in (15), we obtain:

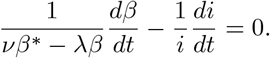

Therefore, for an initial condition (*β*_0_, *i*_0_) at time zero, integration yields:

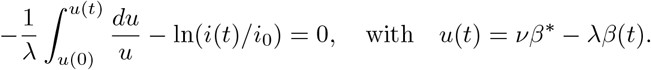

Compressing the logarithms we obtain *λ* ln(*i*(*t*)*/i*_0_) + ln(*u*(*t*)*/u*(0)) = 0, and solving for *u*(*t*) yields *u*(*t*) = *u*(0) · *{i*_0_*/i*(*t*)*}*^*λ*^. Finally,

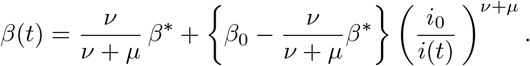

In particular, if the system starts with a transmission rate equal to the intrinsic rate, *i*.*e*., *β*_0_ = *β*^*^, then

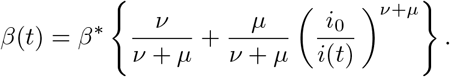

Since *i*(·) is increasing, we have that *β*(·) decreases monotonically.

## 9 Discussion and conclusions

In the context of infectious disease spread processes, our main objective has been the formal analysis, for a group of individuals, of the transition from susceptible to infected epidemiological state. Specifically, our objective was to provide a causal perspective by introducing a novel concept of *contagion force* to explain the rate changes in the transmission dynamics. This work assumes spatial uniformity of the population, an aspect implicit in compartmental models, particularly for the *β*(·)-SI model framework.

This work successfully demonstrates the possibility of establishing – through systemic parameters and variables – constructs analogous to classical mechanics, such as: mass, momentum, and contagion force (of either infectious or susceptible type). This aims to achieve a basic (*i*.*e*., strategic) level of description, understanding, and mechanistic predictability of disease spread phenomena. The goal is to develop a standardized visualization for public health, using broadly accessible mathematical language that facilitates communication and analysis by specialists beyond the research community. We propose a minimal theory of contagion in form, yet one that leverages the intuitive framework provided by physics instrumentation, particularly the kinematics and dynamics of bodies.

**Figure 2.**
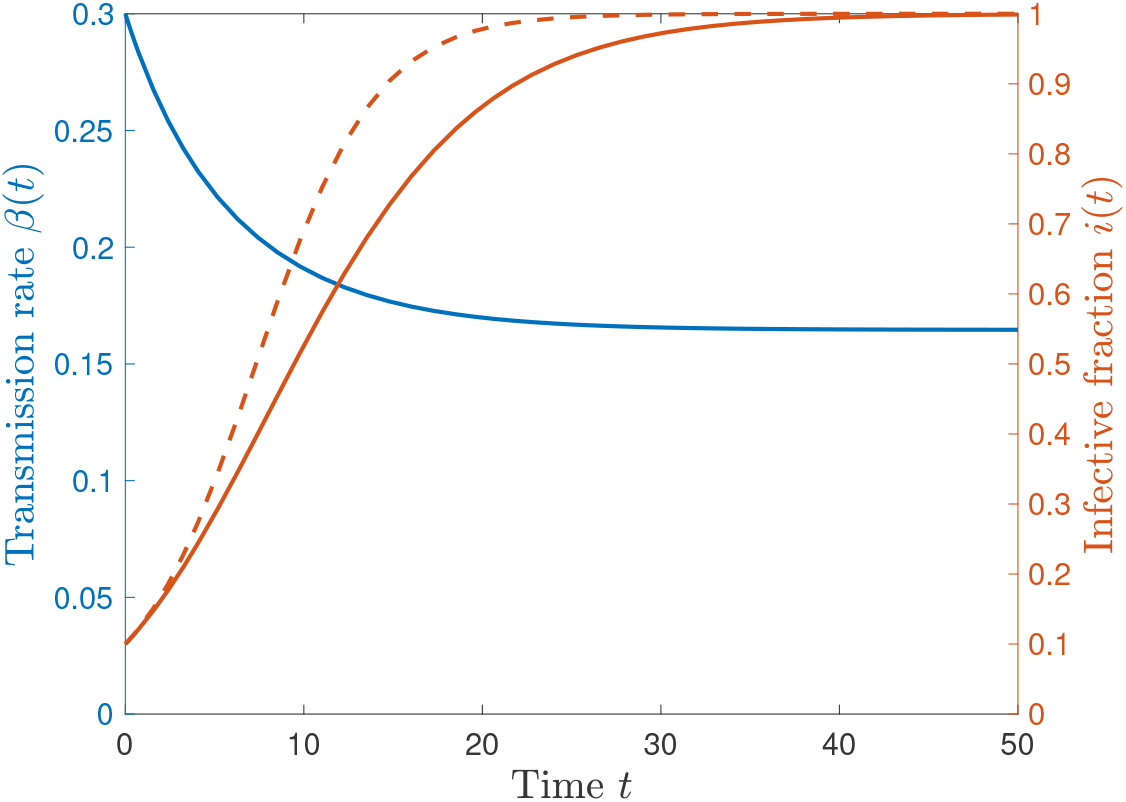
Temporal dynamics of the transmission rate *β*(*t*) and the infective fraction *i*(*t*) according to model (15), with trajectories represented by the blue and red curves, respectively. Furthermore, the red dashed line depicts the trajectory of the classical *SI* model. The parameters are *µ* = 0.3, *ν* = 0.1, and *β*^*^ = *β*_0_ where the initial conditions are *β*_0_ = 0.3 and *i*_0_ = 0.1, from which the long-term value *β*(∞) = 0.1646 is obtained.

Without delving deeper into the broader implications of the introduced mechanical theory, we consider it important to highlight—at least as a plausible hypothesis—how this theory demonstrates linguistic consistency, ultimately expressed through conceptual formulations and their relationships with widely-used mathematics.

Particularly noteworthy is the framing of human (reactive and restorative) behavioral actions as forces that disrupt uniform development conditions. In this regard, a key value of the proposed theory lies in establishing a thinking framework that could serve as a highly useful cognitive-mnemonic model for analyzing contagion phenomena.

In summary, through metaphorical transposition, the theory endows epidemiology with classical mechanics concepts that may already reside in the cultural and educational background of public health specialists—and even lay citizens, who are or should be familiar with basic classical mechanics. Indeed, the epidemiological language framework provided by this *contagion mechanics*, particularly its interpretation of the phenomenon as interplay between forces (especially external forces from human actions or environmental factors), offers greater potential for conceptual clarity. Naturally, this depends on whether the observer already possesses the lens of classical mechanics and, crucially, values the use of such analogies.

The mechanical theory of contagion presented here is explicitly analogic in origin. Its scientific validation will depend on its explanatory power (ex ante) and predictive capacity (ex post). Nevertheless, we argue that the analogy has been rigorously clarified within a well-defined conceptual and mathematical modeling framework.

The proposal will gain strength through deeper empirical validation, *i*.*e*., by demonstrating increasing consistency with real-world data. In this direction, we note that model (15)—apart from its *β*(·)-SEIR context—closely resembles Eq. in (16), which successfully modeled high-resolution daily COVID-19 cases (2020 data) for Chile and Italy (see Figures 7 and 8, respectively, in the cited work). In our view, for simple SIR-type models, this theory exhibits substantial explanatory power.

We note, however, that there remains the possibility that this work constitutes merely a nominalist exercise, failing to incorporate truly novel knowledge—as all analogies break down in one aspect or another, and ours is no exception, (11).

Yet, resolving these doubts will require future developments. For now, we conclude by echoing Bertrand Russell’s words in *Human Knowledge*, Chapter I (61): *“Language serves not only to express thought but to make possible thoughts which could not exist without it*.*”*

## Data Availability

No data used

## Acknowledgments

This research was supported by the Agencia Nacional de Investigación y Desarrollo (ANID), Chile, by the grant FONDECYT Regular No. 1231256.

